# Identifying baseline clinical features of people with COVID-19

**DOI:** 10.1101/2020.05.13.20100271

**Authors:** D. Ferreira-Santos, P. Maranhão, M. Monteiro-Soares, on behalf of COVIDcids

## Abstract

**Objectives:** To describe baseline clinical characteristics of adult patients with COVID-19.

**Methods:** We conducted a scoping review of the evidence available at LitCovid, until March 23th, 2020, and selected articles that reported the prevalence of socio-demographic characteristics, symptoms and co-morbidities in adults with COVID-19.

**Results:** In total, 1 572 publications were published on LitCovid. We have included 56 articles in our analysis, with 89% conducted in China, and 75% contained inpatients. Three studies were conducted in North America and one in Europe. Participants’ age ranged from 28 to 70 years, with balanced gender distribution. Proportion of asymptomatic cases were from 2 to 79%. The most common reported symptoms were fever [4-99%], cough [4-92%], dyspnoea/shortness of breath [1-90%], fatigue 4-89%], myalgia [3-65%], and pharyngalgia [2-61%], while regarding co-morbidities we found cardiovascular disease [1-40%], hypertension [0-40%] and cerebrovascular disease [1-40%]. Such heterogeneity impairs the conduction of meta-analysis.

**Conclusions:** The infection by COVID-19 seems to affect people in a very diverse manner and with different characteristics. With the available data it is not possible to clearly identify those at higher risk of being infected with this condition. Furthermore, the evidence from countries other than China is, at the day, too scarce.

## 1. Introduction

In December 2019, in Wuhan, Hubei Province, China, a cluster of patients with pneumonia of unknown cause was observed [1]. Later, it was found that a new coronavirus caused it. In February 2020, the World Health Organization (WHO) designated the new virus as SARS-Cov-2 and the disease as COVID-19. According to this organization, since the onset of this disease until March 27^th^ 2020, SARS-Cov-2 has infected more than half a million people in 136 countries, leading to the death of 23 335 [2].

The identification of patients that might be infected is crucial, so that they can be adequately screened, treated and/or isolated. In fact, political and health measures have been taken, having in consideration what is supposed to be known about populations at risk (focusing on their baseline co-morbidities) and also identifying those that present a higher chance of being infected by COVID-19 (focusing on their clinical symptoms). However, clinical manifestations are highly variable and the quality of the evidence that underlies these strategies and decisions is frequently not known. We consider that the creation of a predictive model that could help identify those at higher risk of having COVID-19, built on their baseline clinical features (such as sociodemographic, symptoms and presence of co-morbidities), could help prioritize screening and therapeutic strategies. The first step to accomplishing such endeavour is to list the most pertinent variables to be included in such a model. For all this, we have conducted a scoping review to summarize and critically assess articles describing baseline characteristics of individuals infected with COVID-19.

## 2. Methods

### 2.1 Search strategy and selection criteria

To conduct this scoping review, we used the Preferred Reporting Items for Systematic reviews and Meta-Analyses extension for Scoping Reviews (PRISMA-ScR) Checklist [3].

We have reviewed the evidence available on LitCovid [4] for original articles published until March 23^rd^ 2020 in English, French, Italian, Spanish or Portuguese that reported the proportion of socio-demographic characteristics, symptoms and co-morbidities in adults with COVID-19. LitCovid is a curated literature hub for tracking up-to-date scientific information about the 2019 novel Coronavirus indexed and accessible through PubMed. This repository is considered the most comprehensive resource on the subject. We have excluded reviews, opinion articles, case series that included five or less patients, studies that included only pregnant women or children and clear data duplication studies.

### 2.2 Data extraction

Articles were selected by two of the authors independently (DFS and PM) having in consideration the selection criteria. Once the articles were selected, data were extracted (by one of the authors and checked by another) into an Excel spreadsheet and included the following information: date of publication, country of study conduction, method used to detect the presence of COVID-19, last date of participants’ inclusion, type of population, setting, sample size, participants’ age and gender, frequency of asymptomatic patients, and frequency of reported symptoms and comorbidities. We have ordered the included studies by continent, country (by alphabetical order) and sample size (in decreasing order). Only symptoms and co-morbidities described by five or more studies were included in our tables. Those addressed by less than five studies were only described in the narrative synthesis.

## 3. Results

### 3.1 Characterization of the included studies

Until the defined date, there were 1 572 publications in LitCovid and 53 (3%) fulfilled the inclusion criteria. In total, 895 were opinion articles (57%), 50 (3%) had five or less participants included, and the remaining addressed other topics such as diagnostic or genetics. We have used the reference list of 46 (3%) retrieved review articles that had information on frequency of symptoms to identify new articles that were not included in LitCovid database. This procedure led to the inclusion of three additional references [5–7]. In total, we have included 56 studies, as we can see in Figure 1.

**Figure 1.**
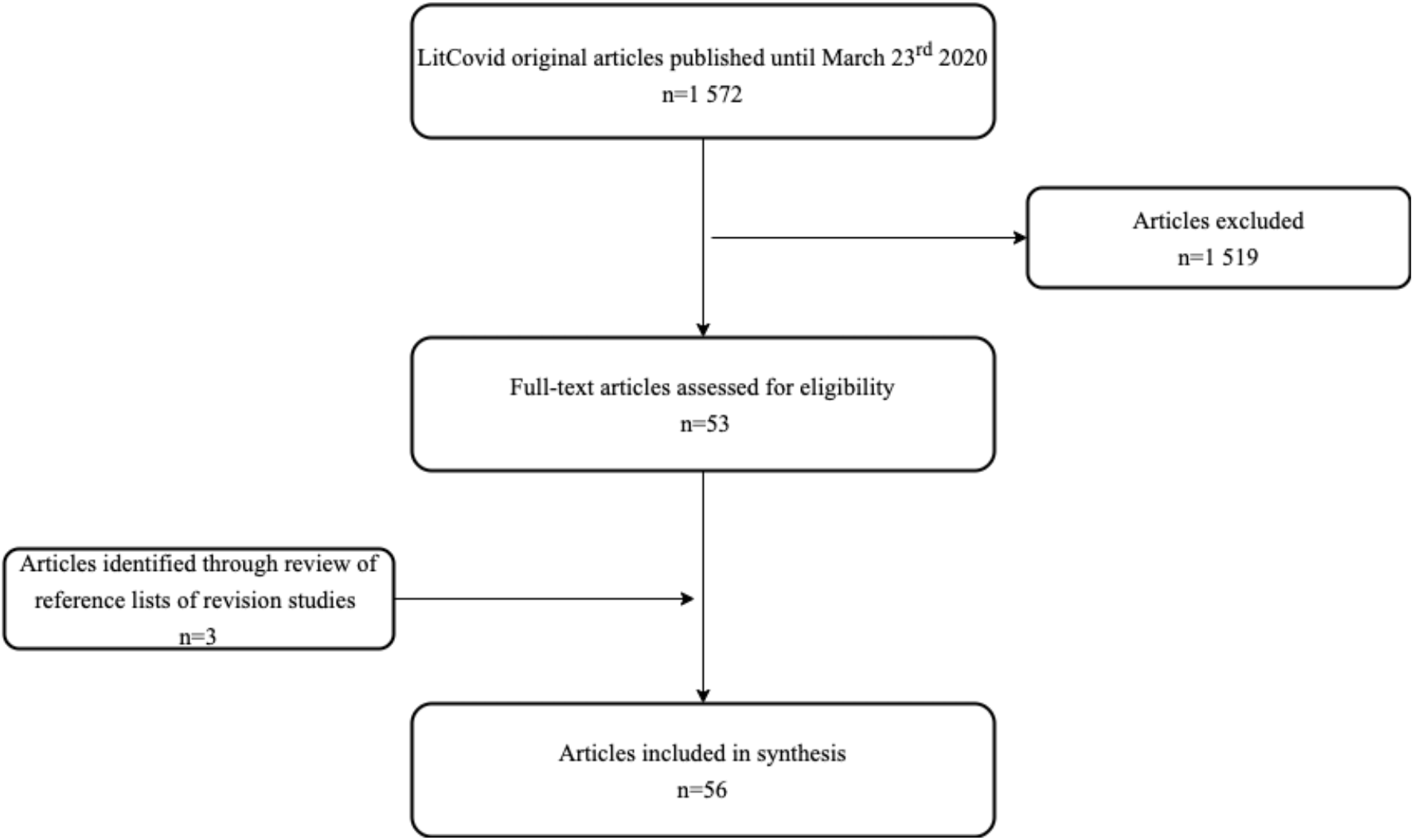
Articles’ selection flow diagram

In Table 1, we can see that, from the included studies, 50 (89%) were from China. We were able to identify only two studies from United States of America (USA), one from Canada, one from Korea, one from Singapore and one multicentre study that included patients from Belgium, Finland, France, Germany, Italy, Russia, Spain and Sweden. No study from Africa or Australia was retrieved.

**Table 1.**
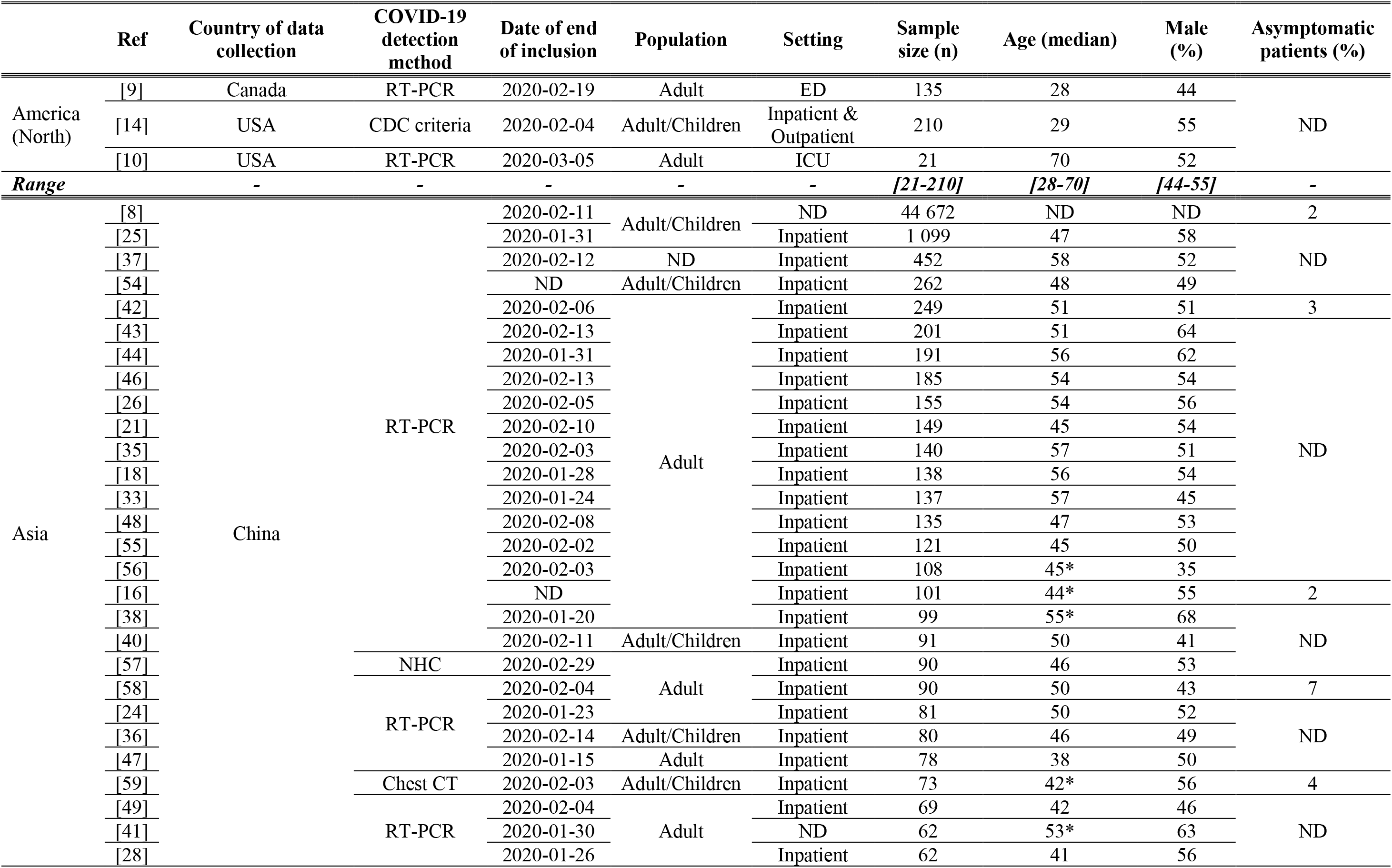
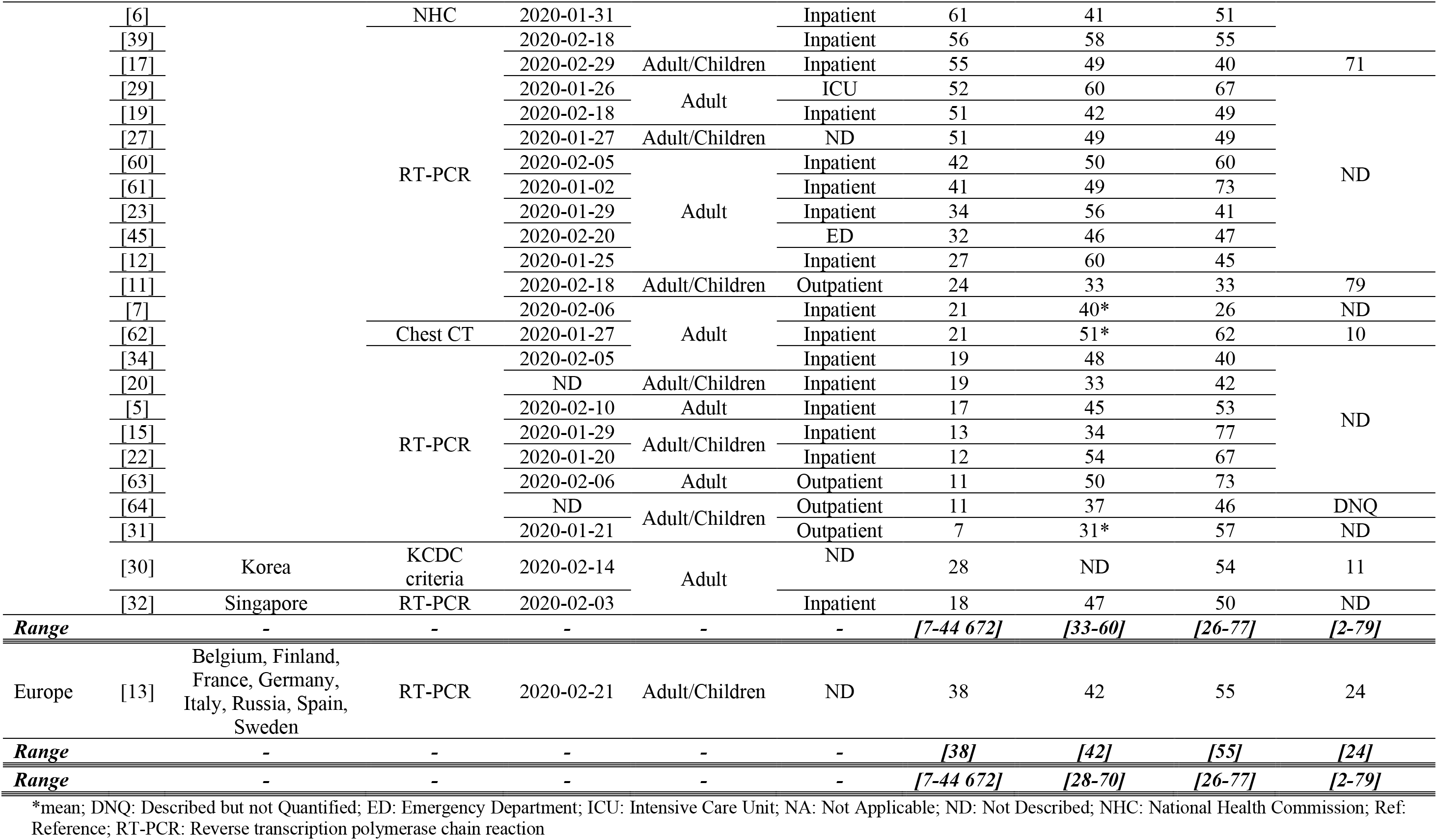
Characterization of the included studies [ordered by continent, country (by alphabetical order) and sample size (in decreasing order)]

The reverse transcription-polymerase chain reaction (RT-PCR) was the method most commonly used to detect the presence of infection by COVID-19 (89%). Looking at the 51 studies that reported setting, two included patients that were seen in the Emergency department, two admitted patients into Intensive Care Unit due to COVID-19, four studies reported included outpatients, one incorporated outpatients and inpatients, while remaining (75%) included inpatients.

In total, 50 500 participants were included. However, one of the studies from China [8] contributed with 88% of the participants. Sample size ranged from 7 to 44 672 participants, with a median of 66 participants per study.

The median age ranged from 28 [9] to 70 [10]. Both studies were conducted in North America. When looking at studies from other continents, we observe a smaller range. In Asia, median age varied from 33 [11] to 60 [12], and 42 in the European multicentre study [13]. There was a balance on gender distribution, with the male gender proportion ranging from 44 [9] to 55% [14] in North American studies, 26 [7] to 77% [15] in Asian studies and 55% in the European study [13]. In 57% of the studies, male gender was more prevalent.

Asymptomatic cases were reported in 10 studies (18%), with no available data for North America. In the European study [13], there were 24% of asymptomatic patients and in Asia it fluctuated from two [8, 16] to 79% [11].

### 3.2 Symptoms

The described symptoms were generally nonspecific and widely variable, ranging from asymptomatic to a rapid multi organ dysfunction as we can see from Tables 2 to 4.

**Table 2.**
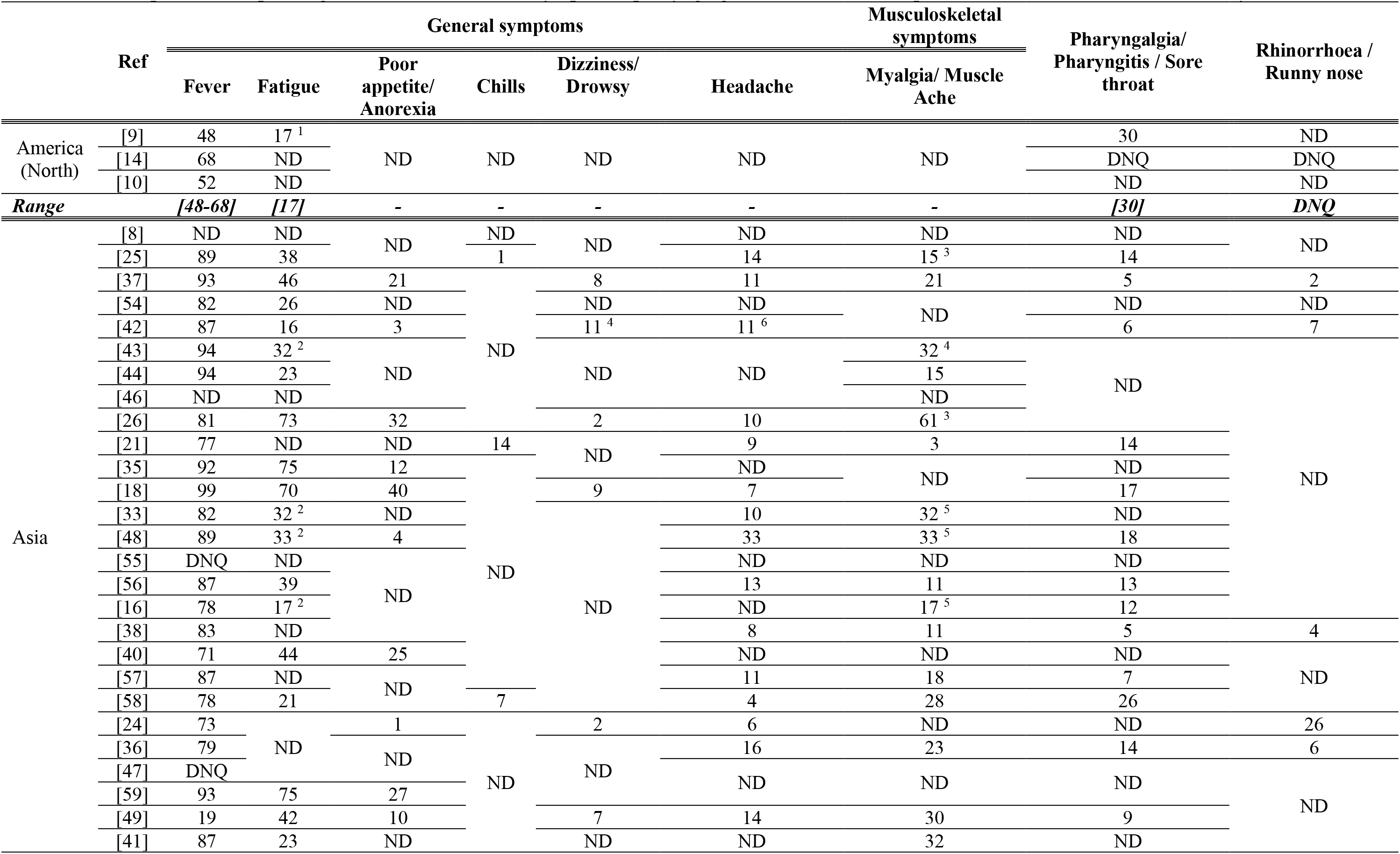
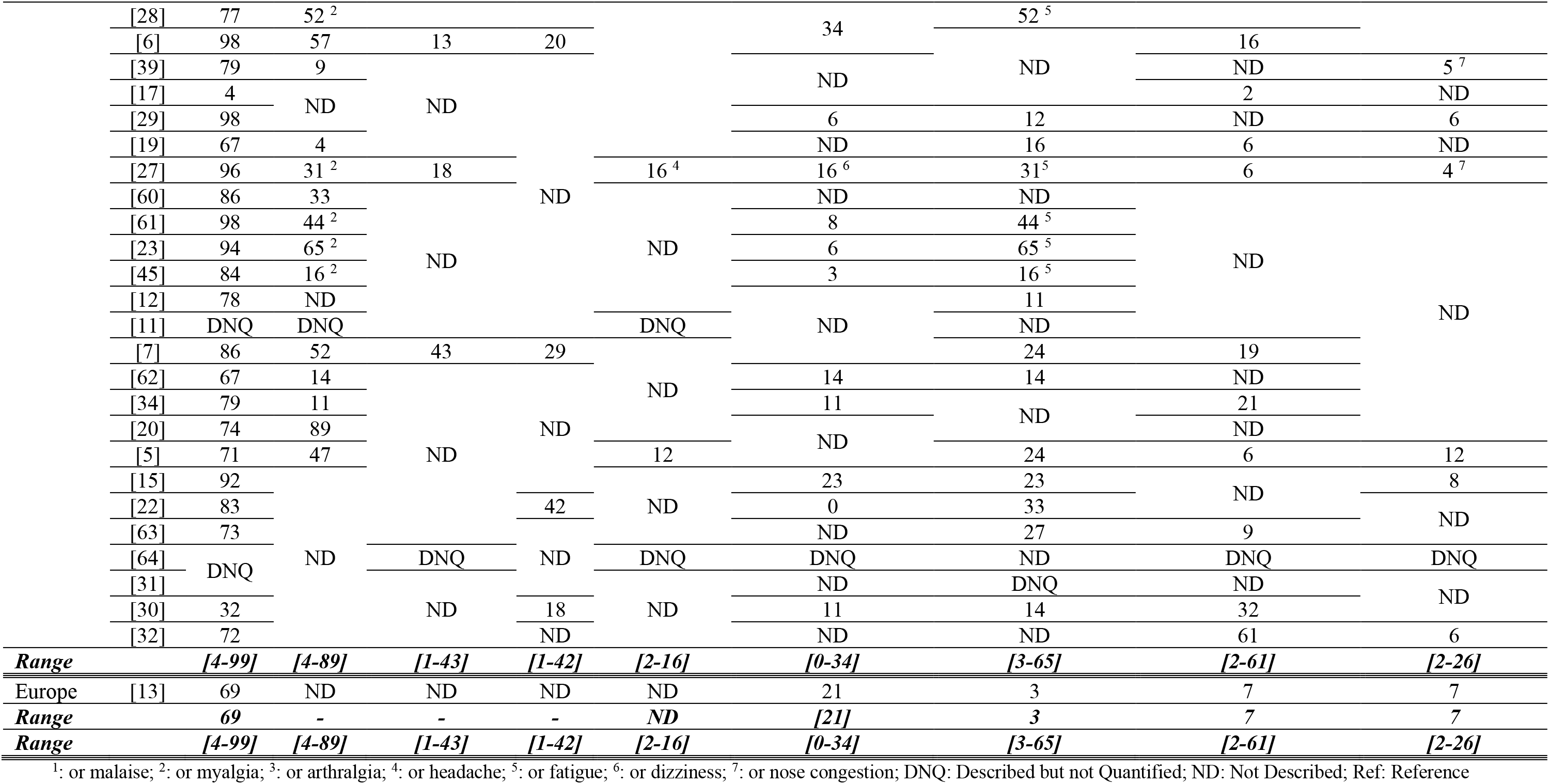
Proportion of reported general, musculoskeletal symptoms, pharyngalgia and rhinorrhoea in patients with COVID-19 at baseline by continent.

**Table 3.**
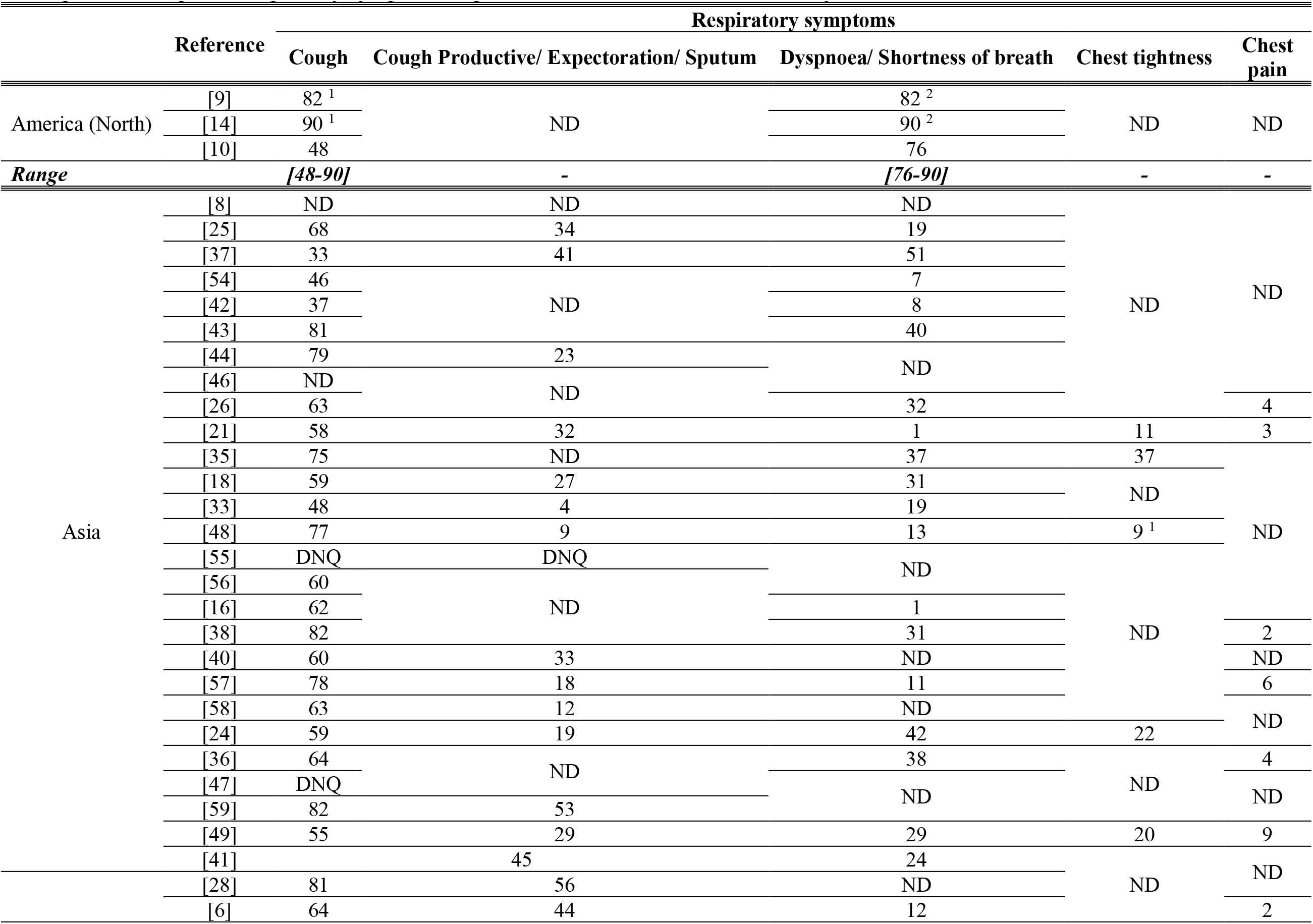
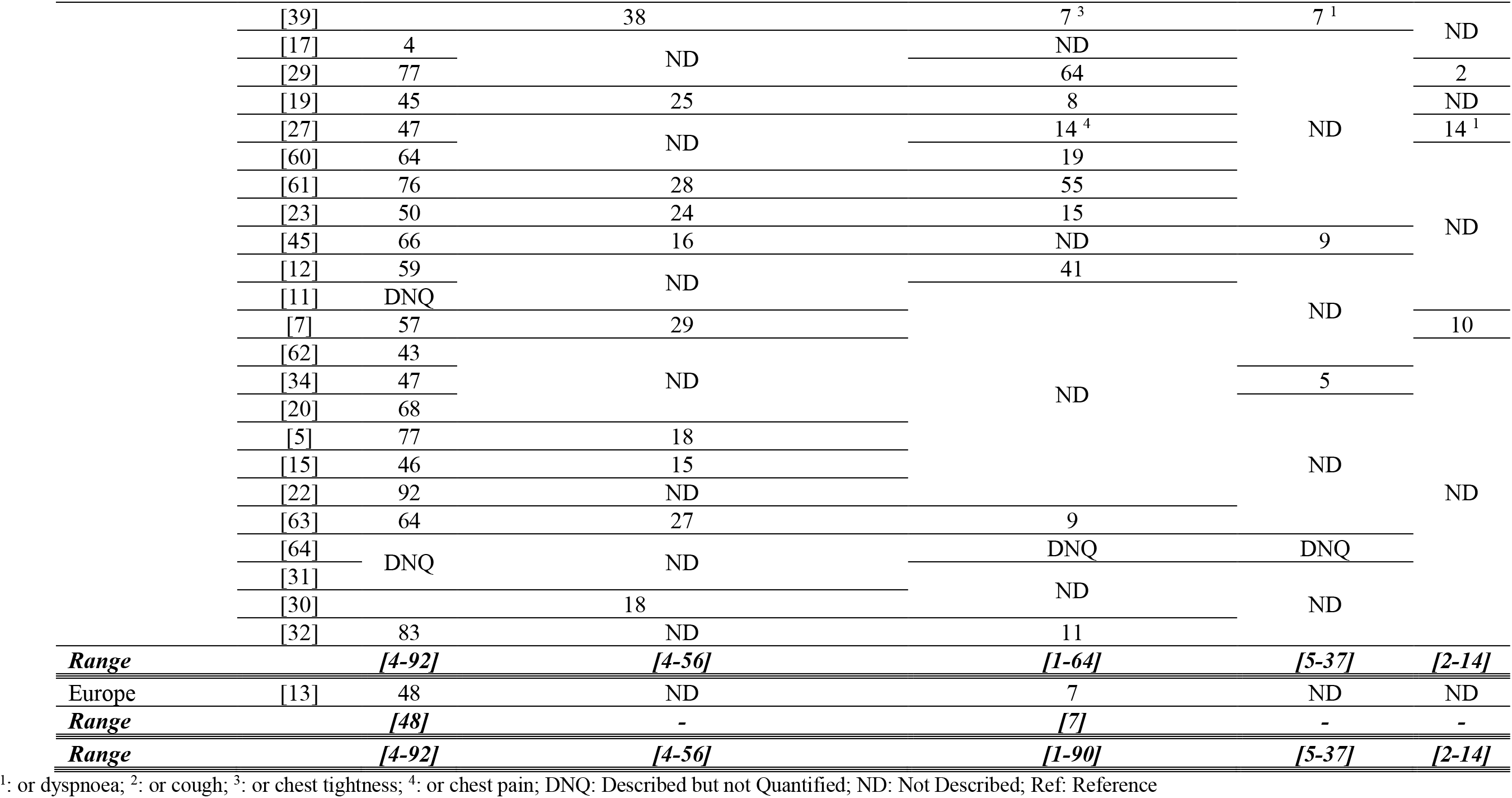
Proportion of reported respiratory symptoms in patients with COVID-19 at baseline by continent.

**Table 4.**
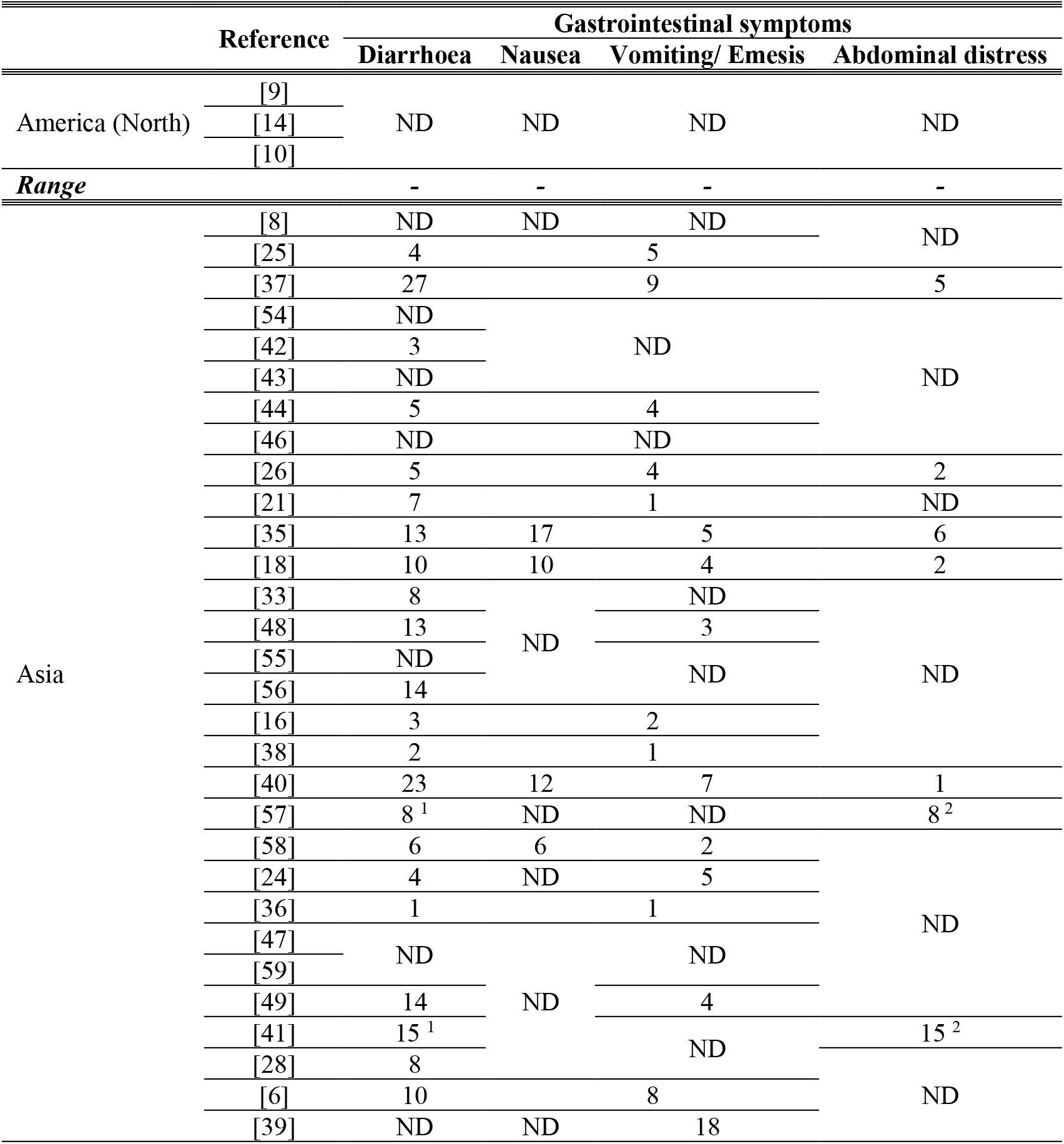
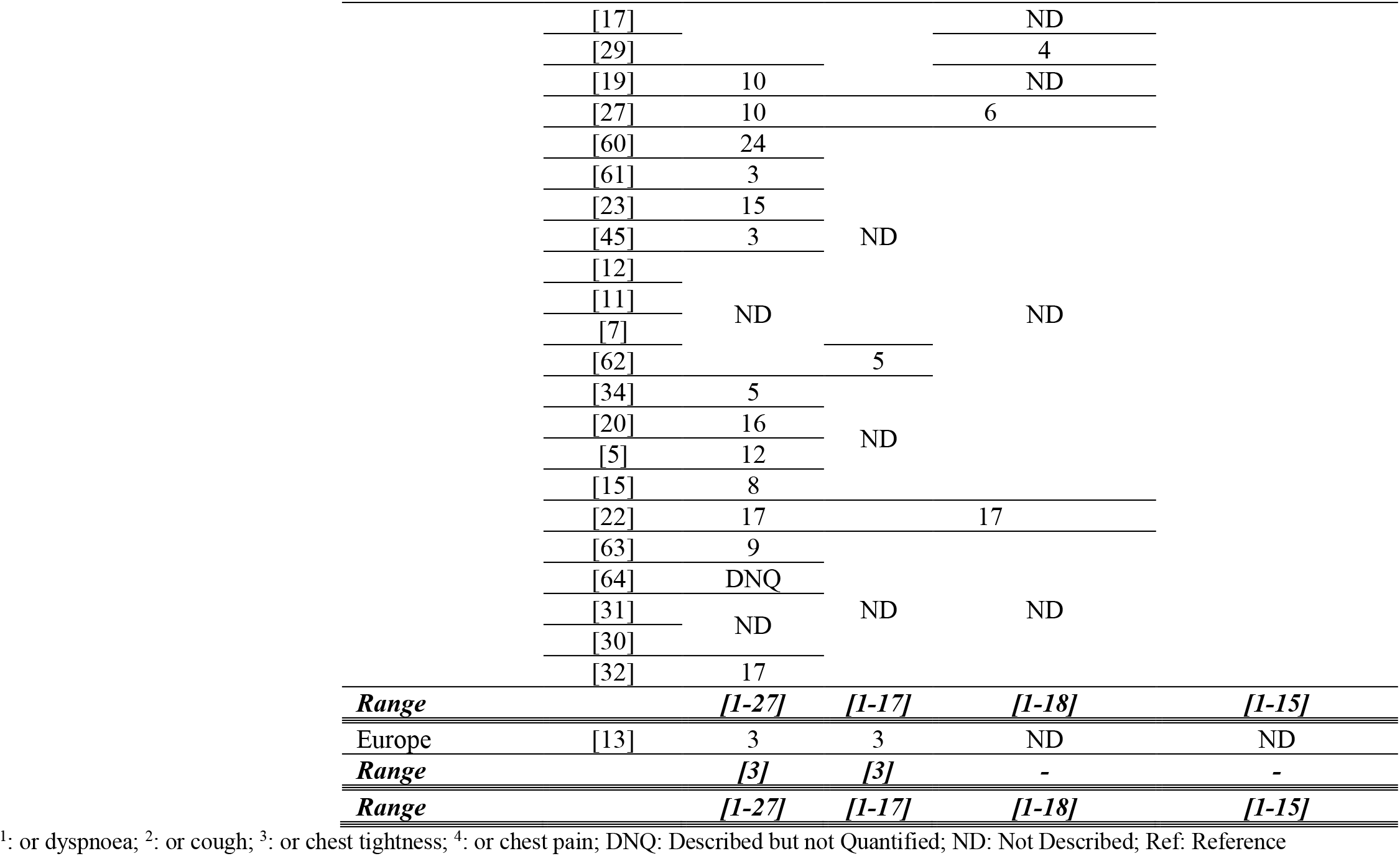
Proportion of reported gastrointestinal symptoms in patients with COVID-19 at baseline by continent.

Fever was one of the most reported symptoms. Its presence ranged from 48 [9] to 68% [14] in North America, 4 [17] to 99% [18] in Asia and 69% in the European study [13].

Fatigue was observed in 17% of the participants from the USA study [14] and ranged from four [19] to 89% [20] in studies from China. Myalgia was reported in 3% of the patients included in the European study [13]. In Chinese studies, it ranged from three [21] to 33% [22], and reached 65% [23] when combined with fatigue. Anorexia, chills and dizziness were registered only in Asian studies and their prevalence ranged from one [24] to 43% [7], one [25] to 42% [22] and two [24, 26] to 16% [27], respectively. Complaints of headache were described in the European study in 21% [13] of the patients and from zero [22] to 34% [6, 28] in two Chinese studies.

Malaise was present in 17% of the participants in one study from USA [9] and 35% in another study from Asia [29]. Weakness was reported in 28% of patients in the European study [13] and ranged from 9 [24] to 11% [30] in two studies from China. Malnutrition was present in two percent of the participants in one study [29]. Skin tingling was described but not quantified in one study [31]. Arthralgia was described in three studies, all conducted in China [25, 26, 29]. This symptom was reported in two percent of the sample in one study [29], and 15% [25] and 61% [26] in two studies that combined the presence of arthralgia or myalgia as one symptom.

The presence of pharyngalgia was reported in one study from USA [14], in 30% of the participants.. In studies from China the prevalence of this symptom varied from two [29] to 61% [32]. In the European study [13], both pharyngalgia and rhinorrhoea were reported by 7% of the participants. The later symptom ranged from two [29] to 26% [24] in Chinese patients. The frequency of nasal congestion and throat congestion was reported only in studies from China. In one study [25], two percent of the participants described feeling throat congestion, and nasal congestion varied from 5 [25] to 62% [15] in two studies.

From respiratory symptoms, cough was the most frequently assessed; one study from USA and European study reported to be present in 48% of the sample [10, 13]. Cough or dyspnoea were reported by 82% of the patients in one study from Canada [9] and 90% in the USA [14]. Specifically, productive cough, chest tightness and chest pain were registered only on studies from China and varied from 4 [33] to 56% [28], 5 [34] to 37% [35], and two [29] to 14% [27], respectively. In one study [27], 14% of patients reported feeling chest pain or dyspnoea.

The presence of dyspnoea alone was also described in the majority of the studies. One study from the USA reported its presence in 76% of the patients [10], while in the European study [13] it was only observed in 7% of the patients. As for the studies conducted in China, dyspnoea prevalence oscillated from one [21] to 64% [29].

General gastrointestinal symptoms were described by 10% [9] of the patients in one study from Canada and 40% [35] in another from China. From the gastrointestinal system, only diarrhoea and nausea were recorded in the European study [13]. Both presented a 3% prevalence. From the studies conducted in China, diarrhoea prevalence ranged from one [36] to 27% [37], nausea from one [21, 36, 38] to 17% [22], vomit from one [29] to 18% [39] and abdominal distress from one [40] to 6% [35]. When combining abdominal pain or diarrhoea, the prevalence raised to 15% [41]. Belching or gastritis was recorded in only one study from China [35] and was reported by 5% of the patients. Irritability or confusion was documented in three [21] and 9% [38] of the patients included in two studies from China, and the presence of rash and enlargement of lymph nodes was assessed in only one study and was not found in any patient [25].

### 3.3 Co-morbidities

As we can see in Table 5, the presence of co-morbidities was not reported in the European study and only one of the studies from the USA had relevant information [10]. In this study, 86% of the patients had at least one comorbidity. The most frequent were chronic kidney disease (48%), congestive heart failure (43%), diabetes (33%), chronic obstructive pulmonary disease (33%) and obstructive sleep apnoea (29%). Less than 10% of the patients presented end-stage kidney disease, asthma, cirrhosis and rheumatologic disease.

**Table 5.**
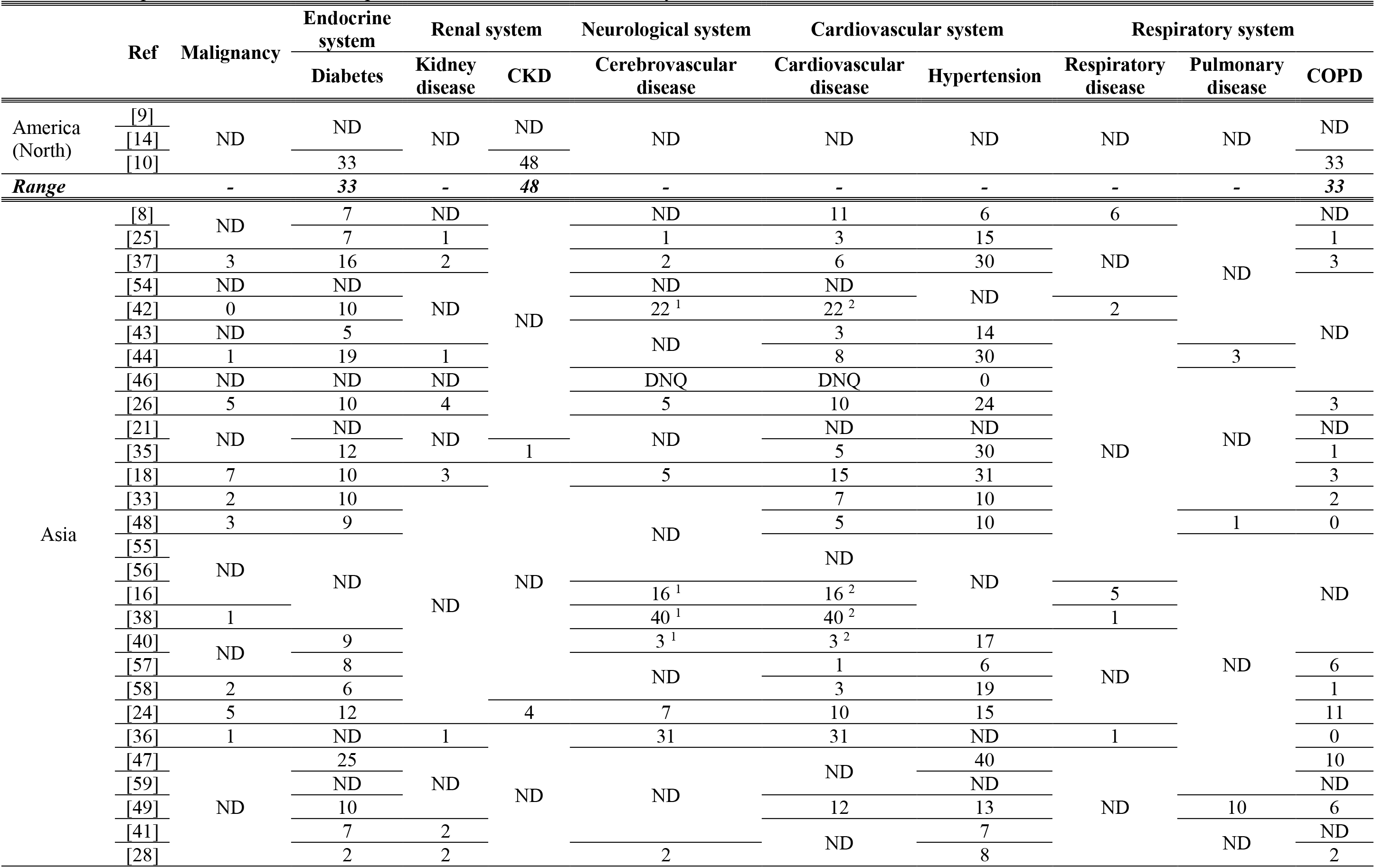
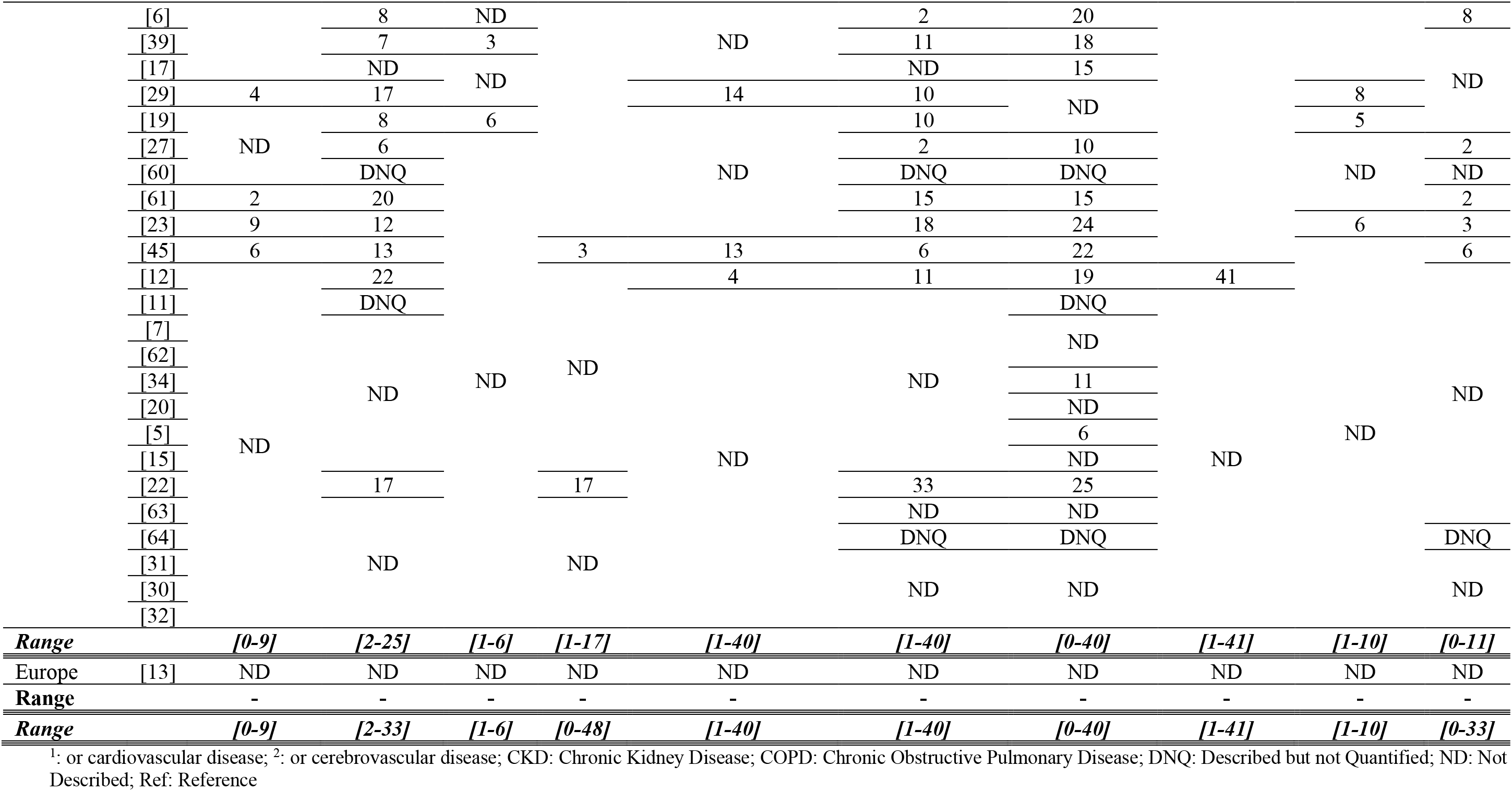
Proportion of co-morbidities in patients with COVID-19 at baseline by continent.

The remaining data was from Asian studies, in which several concomitant infections were described. The presence of Hepatitis B was observed in one [42], two [17, 25] and 5% [34] of the participants in the four studies that described its frequency. Prevalence of human immunodeficiency virus was reported to be of zero [18, 26, 36] and 6% [23]. Only one study described bacterial co-infection in 17% of the patients [22].

Numerous studies described that some patients presented malignant diseases. This co-morbidity prevalence ranged from zero [42] up to 9% [23]. Only one study described the presence of thyroid disease [35] and other of hyperlipidaemia in four and 5% [35] of the participants, respectively. Two studies reported the presence of hypothyroidism in two [17] and 6% [23] of the patients. Various studies reported the prevalence of diabetes, with values ranging from 2 [43] up to 33% [44]. The presence of kidney disease ranged from one [8, 25, 44] up to 6% [19]; chronic kidney disease was observed in one [35] up to 17% [24] of the patients. Only one study reported the proportion of patients with renal insufficiency [22] and urolithiasis [35] to be of 17% and two percent, respectively. Chronic liver disease was observed in zero [22] to 11% [28] of the participants. Hepatic insufficiency was reported by two studies to have a prevalence of 9 [24] and 17% [22]. Fatty liver and abnormal liver function were observed in 6% of the patients in one study [35]. Digestive system diseases were described in four studies in four [42], 6 [16] and 11% [38] of the participants.

The presence of cerebrovascular disease was reported in several studies and ranged from one [25] to 31% [36] of the participants, reaching 40% [38] when combined with cardiovascular disease. Dementia was described in one study, with a value of two percent [29]. Nervous system diseases were ascertained in three studies, with a frequency of one [36, 38] and three percent [45]. The same number of studies registered a history of stroke and observed its presence in two [35, 41] and 8% [22] of the participants.

Cardiovascular disease prevalence ranged from one [6, 27] to 33% [22] of the patients in the various studies reporting it. Hypertension frequency varied from zero [46] to 40% [47]. The presence of tachycardia was registered in four studies and reported to be of two [6], four [48] and 7% [33, 49]. We only found one study that described the prevalence of arrhythmia (with a value of four percent [35]), persistent atrial fibrillation (6% [39]), cardiac failure (8% [22]) or aorta sclerosis (1% [35]).

Various studies described the prevalence of baseline respiratory system conditions. Respiratory disease, in general, was found in one [36, 38] to 41% [12] of the patients, pulmonary disease to range between one [48] to 10% [49], and chronic obstructive pulmonary disease between zero (37, 47) and 33% [49] of the patients. We only found two studies that described the prevalence of asthma (with a value of two percent [17] up to9% [10]), and one study describing a 6% of rhinitis [5].

## 4. Discussion

This is the first scoping review focusing on baseline characteristics of patients with COVID-19. Although our aim was to try to better identify those at higher risk of having the condition, only descriptive studies were found. We have identified 56 articles; two were conducted in USA, one in Canada and one was a multicentre European study. No studies from Africa, South America or Australia were retrieved. At the date of the end of our review, according to WHO [50], there were 25 375 cases of COVID-19 in the Region of the Americas, 171 424 in European Region and 990 in African Region.

As we can observe above, most of the studies were conducted in China, the first country in which COVID-19 was detected. Furthermore, one of these studies [8] contributed to 88% of the participants. This study consists of the Chinese Center for Disease Control and Prevention Report.

Therefore, we cannot be sure about how many of the other Chinese studies described results that are included in this report, representing duplicate participants.

We observed a very high heterogeneity on sample size, patients’ age and described symptoms and co-morbidities. Accounting for this heterogeneity, we have considered that it was not adequate to conduct a meta-analysis and performed only a narrative synthesis of the available evidence. We also acknowledge the exclusion of articles written only in Chinese due to the fear of further data duplication [51].

In the included studies, the median age ranged from 28 to 70 years, being 50 years or less in 36 (72%) of the studies. Only one-fifth of the studies described the proportion of asymptomatic patients. In the European study, it was around 25% and in the Asian studies ranged from two up to 75% of the patients. It highlights the importance of wide screening and people isolation strategies due to the risk of being in contact with infected but asymptomatic people.

The prevalence of more than 30 symptoms and 35 co-morbidities were collected; however, several were reported by five or less studies. The most reported symptoms were fever, cough, dyspnoea, fatigue, myalgia and pharyngalgia. Cardiovascular disease, hypertension and cerebrovascular disease were the most reported comorbidities. However, this is also due to the commonly high prevalence of these diseases in the general population and the focus given to more severe cases by several studies.

There is a previous systematic review with meta-analysis of prevalence of symptoms and comorbidities in people with COVID-19 that included eight studies published until February 5, 2020 [52]. The authors concluded that the most prevalent clinical symptoms were fever (with a pooled prevalence of 91%), cough (67%), fatigue (51%) and dyspnoea (30%). The most prevalent comorbidities were hypertension (17%), diabetes (8%), cardiovascular diseases (5%) and respiratory system diseases (2%). However, the authors reported high levels of heterogeneity when pooling such prevalence (I^2^ ranged from 85 to 96%).

A more recent systematic review with meta-analysis to identify clinical, laboratory and imaging features of COVID-19, included studies until February 21, 2020 [53]. When pooling the 18 included studies, once again, fever (pooled prevalence of 88%), cough (58%) and dyspnoea were the most common symptoms, and hypertension (19%), cardiovascular disease (14%) and diabetes (12%) the most frequent co-morbidities. Once again, severe heterogeneity was observed by the authors.

Our study, observed that the presence of fever ranged from four to 99%, cough from four to 92%, fatigue from four to 89% and dyspnoea from one to 90%; as for co-morbidities the prevalence of hypertension varied from zero to 40%, diabetes from 2 to 33% and cardiovascular disease from one to 40%. We highlight that these values cannot be directly compared between studies without having in consideration that they reflect the existence of different populations, healthcare settings, selection criteria and different times of the disease history. Such massive variation on the range of observed prevalence for all symptoms and co-morbidities impairs the selection of any of them as pertinent to be included in a predictive model to identify people at high risk of being infected with COVID-19.

We consider that future research conducted specifically with that aim and assessing the ability of several symptoms and/or co-morbidities combined to stratify people by their risk of being infected is crucial. Also, there is a great need for further studies conducted outside China so that comparisons can be made about baseline characteristics as well as clinical outcomes.

## Data Availability

All data is described in LitCOVID (https://www.ncbi.nlm.nih.gov/research/coronavirus/)

## Acknowledgments

The work from Daniela Ferreira-Santos was supported by Fundacão para a Ciência e Tecnologia (FCT) [grant number PD/BD/13553/2018]. The work from Priscila Maranhão was supported by ODISSEIA – Oncology Information System project (POCI-05-5762-FSE-039021), financed by the North Portugal Regional Operational Programme (NORTE 2020), under the PORTUGAL 2020 Partnership Agreement, and through the European Regional Development Fund (ERDF) and European Social Fund (ESF) respectively. The authors declare that they have no conflicts of interest. We also would like to acknowledge Professor Pedro Pereira Rodrigues and M.D. Ana Margarida Pereira, for a critical review of the manuscript.

